# The Effectiveness of Advance Care Planning Training for Care Home Staff: a Systematic Review

**DOI:** 10.1101/2023.02.28.23286494

**Authors:** Victoria Ann Barber-Fleming, Mala Mann, Gillian Mead, Aoife Gleeson

## Abstract

In line with population ageing, the number of global deaths is predicted to increase. There have been projections that, within the next 20 years, in England and Wales, care homes may become the most common place of death. In order to respect the autonomy of their residents, it is therefore, vital that care home staff are able to have Advance Care Planning conversations. However, care home staff may lack the knowledge or confidence to have such discussions. Further, a systematic review found a paucity of evidence about whether Advance Care Planning training interventions for care home staff are effective. New, higher quality studies are now available, justifying this review update. We sought to address two questions: 1) ‘What Advance Care Planning education interventions exist for care home staff?’ and 2) ‘how effective are these interventions?’ All measurable outcomes of effectiveness (e.g. health system/resource-related, patient/relative-related, staff-related) including both qualitative and quantitative measures of effectiveness were considered.

**Design:** The review adheres to the Preferred Reporting Items for Systematic Reviews and Meta-Analyses (PRISMA) and is registered on PROSPERO (ID: CRD42022337865). Original research evaluating Advance Care Planning education for care home staff and reporting any measurable outcome of effectiveness was included. We searched Ovid Medline All, Ovid Embase, Cochrane Central Register of Controlled Trials, EBSCO CINAHL, EBSCO ERIC, and Ovid PsycINFO from March 2018 (3 months prior to original review search cut-off) to June 2022, with supplemental journal and website searches. The results were synthesised by narrative synthesis.

**Findings:** The current review update almost doubled the number of included studies in a relatively short period. This review includes 10 studies (n = 310 care homes), from the UK, Belgium, Norway and Canada. UK studies were mainly related to the Gold Standard Framework for Care Homes. Two studies adopted multi-component education interventions. Outcome measures included resident/family, staff and health service-related concepts. Even after identifying a further 5 papers, there remains insufficient evidence to determine the effectiveness of Advance Care Planning education interventions for care home staff.

**Conclusions:** Advance Care Planning education interventions are heterogeneous and often complex in their design, flexibility, target populations, and outcomes. There remains insufficient data to determine the effectiveness of Advance Care Planning education interventions for care home staff, with a particularly urgent need to agree on outcome measures of the effectiveness. Future research could consider updating the existing Delphi consensus on outcome measures for evaluating Advance Care Planning, in light of this systematically collected evidence, with a view to agreeing outcomes that are specific to Advance Care Planning education interventions for care home staff.

## Introduction

The increasing rate of global population ageing is a success story and grand challenge, demanding that countries prepare their health and social care systems urgently (World Health Organisation, 2022). Globally, the number of people aged over 65 years is predicted to double in the next 30 years (United Nations, 2019). In the UK, the cohort of people aged over 85 years is ageing most rapidly, with predictions that this group will account for 7% of the UK population by 2041 (Office For National Statistics, 2018). Models for England predict that the increase in numbers of ‘older old’ adults will result in this cohort experiencing double the number of people living with low-dependency and almost double living with high dependency (Kingston et al., 2018). This is reflected in predictions that by 2030, the number of care home residents in the United States is expected to double (Kelley & Morrison, 2015) and by 2040, care homes will be the most common place of death in the England and Wales (Bone et al., 2018).

In the UK, ‘care homes’ offer a spectrum of support options, including accommodation, social activities and personal care, and ‘nursing homes’ offer additional nursing care, with some homes able to offer care specifically for those living with dementia (Age UK, 2022). Terminology differs globally, therefore, for the purposes of this global review, the terms care home, nursing home and long-term care will be synonymous. People are entering care homes later in their frailty trajectory (British Geriatric Society, 2021), resulting in over 50% of U.S care home residents requiring assistance with most activities of daily living (Kelley & Morrison, 2015) and up to 75% of older adults in UK care homes living with dementia (British Geriatric Society, 2021). With up to 56% of residents dying within a year of admission (Kinley et al., 2014a), it is perhaps unsurprising that care homes are now referred to as “the defacto hospice” (Johnston et al., 2022).

This increasing, complex morbidity and mortality in care homes highlights the importance of care home staff being prepared to manage the palliative care needs of their residents. A related skill, required of care home staff, is to support Advance Care Planning (ACP) discussions with residents and families. International consensus is that ACP can be defined as “enables individuals to define goals and preferences for future medical treatment and care, to discuss these goals and preferences with family and health-care providers, and to record and review these preferences if appropriate” (Rietjens et al., 2017). An overview of 80 systematic reviews of ACP interventions found that interventions and outcome measures are diverse and the quality of studies in the field is poor (Jimenez et al., 2018). The authors found evidence that ACP may improve communication, discussion and documentation of end of life choices, the likelihood of dying in ones preferred place of care, and have cost savings for the health system. Specific to ACP interventions in care homes, a systematic review of 13 studies found that they were most commonly evaluated in respect to hospitalisations, which were reduced by 9-26% (Martin et al., 2016). Further, the interventions were associated with 29-40 % more people dying in the care home and 13–29% increase in alignment between resident’s wishes and actual experiences. The ACP process is extremely relevant to care home residents as they commonly follow a “dwindling” trajectory of decline, associated with frailty, which can be difficult to recognise and forecast (Kinley et al., 2014a). Unexpected deteriorations in the care home resident’s medical condition, often results in unplanned hospital admission (Spacey et al., 2018). Further, residents with dementia are uniquely exposed to violations of their autonomy as they lose their ability to communicate their preferences, making it especially important to have timely ACP conversations (Flo et al., 2016). Despite older adults being willing to engage in ACP discussions (Sharp et al., 2013; Mignani et al., 2017) they do not happen frequently enough in the care home setting (Mignani et al., 2017).

Further, the COVID-19 pandemic disproportionately impacted those living and working in care homes. A rapid review (Selman, 2020) including 21 primary studies and 10 systematic reviews found that the pandemic has both facilitated and hindered progress in developing ACP in the community. Positive impacts include improving public awareness and technology around the process, and negative impacts include worsening national coordination of resources and systems.

Molloy et al. (2000) suggest that residents will become involved with ACP if they are offered the opportunity to do so. However, not all care home staff have the appropriate knowledge of basic end of life care management (Smets et al., 2018) and staff feel unsupported in managing palliative care issues (Macgregor et al., 2021). This lack of knowledge and self-efficacy (Gilissen et al., 2020) results in staff evading ACP discussions (Spacey et al., 2018). Qualitative research has shown that care home staff are supportive of the ACP process in principle, but underprepared for these discussions and frustrated when resident’s wishes are not aligned with the actual care they receive (Vellani et al., 2022). Education of care home staff is therefore a priority to improve staff knowledge and self- efficacy in ACP.

Palliative care education interventions for care home staff are highly variable in their methods (Lamppu & Pitkala, 2021). Specifically, in relation to ACP education interventions in care homes, Gleeson et al. (2021) found that only 6 studies met their inclusion criteria and they were heterogeneous in size, method and quality. This limits the ability to synthesise research findings into a meaningful, evidence-based approach to ACP education interventions for care homes. A review update is the best approach to answer the review questions as 1) they remain extremely relevant, with strategic importance for clinical practice and national guidelines, perhaps even more so in light of the pandemic impact, and 2) there is new, high quality evidence which may impact the review findings and improve credibility of the well-designed original review (Garner et al., 2016). The un-standardised approach to educating care home staff, combined with the unique challenges faced by care homes in relation to high staff turnover and diversity in experience and knowledge of staff, renders ongoing review of education interventions urgent (Iida et al., 2021).

## Aims

This review aims to address the continuing gap in the literature about the effectiveness of ACP interventions to educate all levels of health care professionals working in care homes. Our questions are 1) ‘What anticipatory care planning education/training interventions exist for care home staff?’ and 2) ‘how effective are these interventions?’

## Methods

### Search Strategy

The review adhered to the Preferred Reporting Items for Systematic Review and Meta-Analysis (PRISMA) 2020 guidance (Page et al., 2021). Gleeson et al. (2021) searched from inception to June 2018. Their search strategy was replicated, with modifications to subject headings as appropriate in relation to updates. To compensate for time-lags in publication, the updated search included March 2018 to June 2022. The recorded ‘create date’ rather than ‘publication date’ was used, where possible within the databases, to find eligible studies. The search was conducted on 14^th^ June 2022 in Ovid Medline (R), Ovid Medline (R) and Epub Ahead of Print, In-Process, In-Data-Review & Other Non-Indexed Citations, Daily and Versions, Ovid Embase, Cochrane Central Register of Controlled Trials (CENTRAL), EBSCO CINAHL, EBSCO ERIC, and Ovid PsycINFO. Supplementary searches were conducted on 20^th^ June 2022 in websites (The National Council for Palliative Care, Association for Palliative Medicine of Great Britain and Ireland, Ehospice, European Association for Palliative Care, Gold Standards Framework) and journals (American Journal of Hospice and Palliative Medicine, BMJ Supportive and Palliative care, Journal of Hospice and Palliative Nursing, International Journal of Palliative Nursing and Palliative Medicine). The full search strategy for each database is available in Supplementary File 1. We registered the protocol on the International Prospective Register of Systematic Reviews (PROSPERO Registration: CRD42022337865).

### Population, Intervention, Comparison, Outcomes, Study design (PICOS): Inclusion and Exclusion Criteria

- *Population*: any staff working within a care home. Note this is amended from the original search and protocol which specifies ‘health care professionals’ as the population (Gleeson et al., 2021). The change reflects the multi-disciplinary nature of care home teams (including social care and administration staff).
- *Intervention:* ACP education or training for care home staff, where ACP training is the overarching focus of the intervention and ACP interventions are for care home staff (not patients or families) and for use within care homes (not hospital or hospice settings). Exclude ACP education interventions which focus on a specific disease (e.g. cardiac disease) other than dementia. Note, this is justified by the high number of care home residents who live with dementia.
- *Comparison:* may include no intervention/usual care or alternative intervention or comparison within groups in before and after studies.
- *Outcomes:* all measurable outcomes of effectiveness (e.g. health system/resource-related, patient/relative-related, staff-related) including both qualitative and quantitative measures of effectiveness.
- *Study design:* studies in English language with full text available via University databases. Original research studies with quantitative, qualitative or mixed-methods designs will be considered for inclusion. Studies must have a measurable outcome in relation to effectiveness of the ACP education intervention.

### Study Selection

Searches of the above databases, websites and journals were carried out by VBF. EndNote Library was used to batch the exported studies and de-duplication was conducted using the Systematic Review Accelerator De-duplicator tool (Clark et al., 2020), with subsequent manual de-duplication of any remaining duplicates. Title and abstract screening and full text screening were conducted in Covidence, by VBF and AG, who independently selected studies for inclusion, against the predetermined inclusion criteria (PICOS). Any disagreements were resolved by discussion between the two reviewers. The screening process and number of included studies is described in the PRISMA Flow diagram (Figure 1).

**Figure 1.**
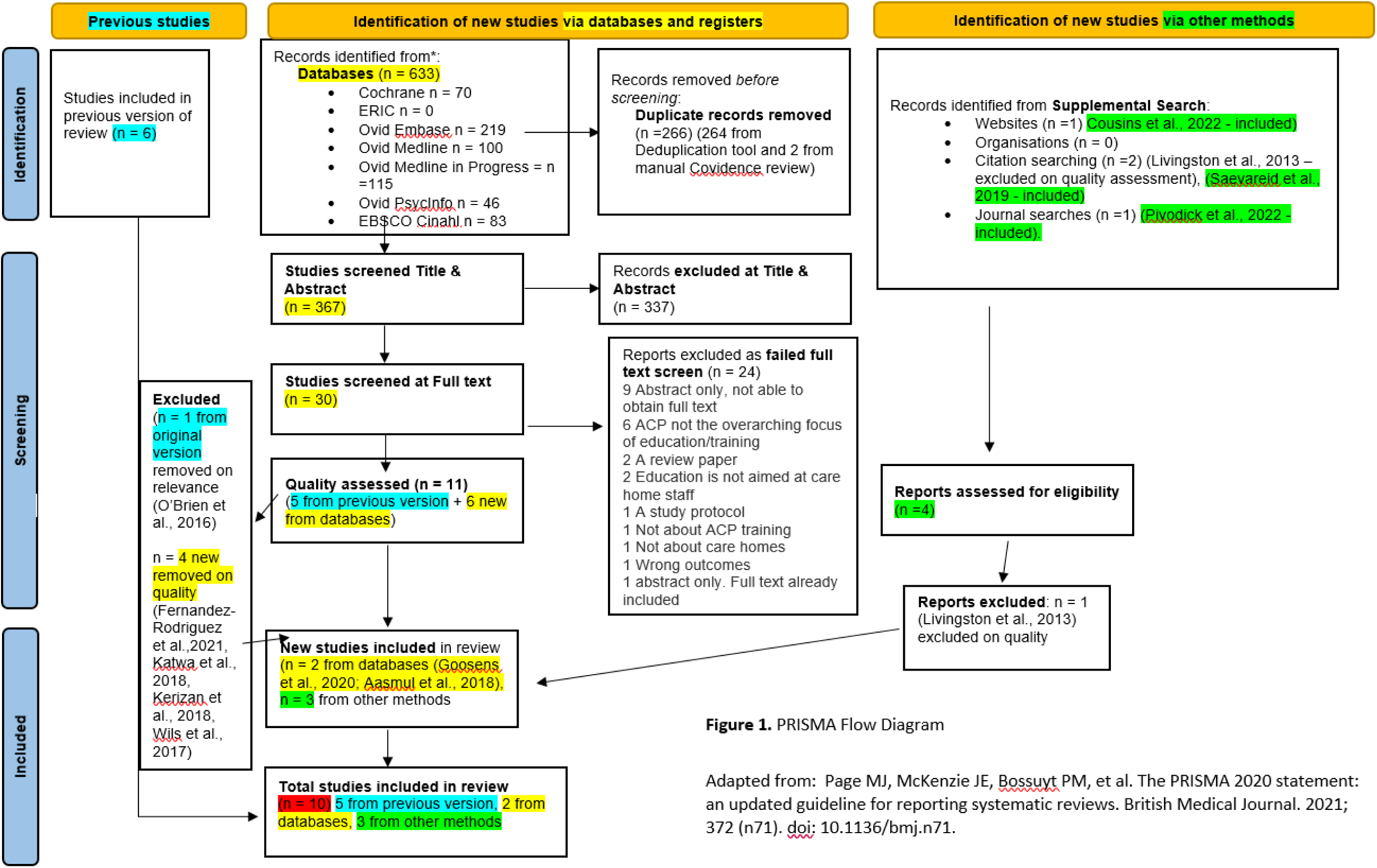
Prisma Flow Diagram

### Data Extraction

VBF and AG initially piloted data extraction from 5 studies using the extraction form from the original review, allowing us to sense check and align our data extraction processes. VBF then independently extracted the following data: study design, aim, setting, participants, inclusion and exclusion criteria, intervention design, allocation method, control, sample size, outcome measures, results and ethical approval. Table 1 presents the characteristics of included studies. Table 2 groups the intervention design and outcome measures by country. Table 3 provides more detail about the staff participants in the primary studies.

**Table 1:**
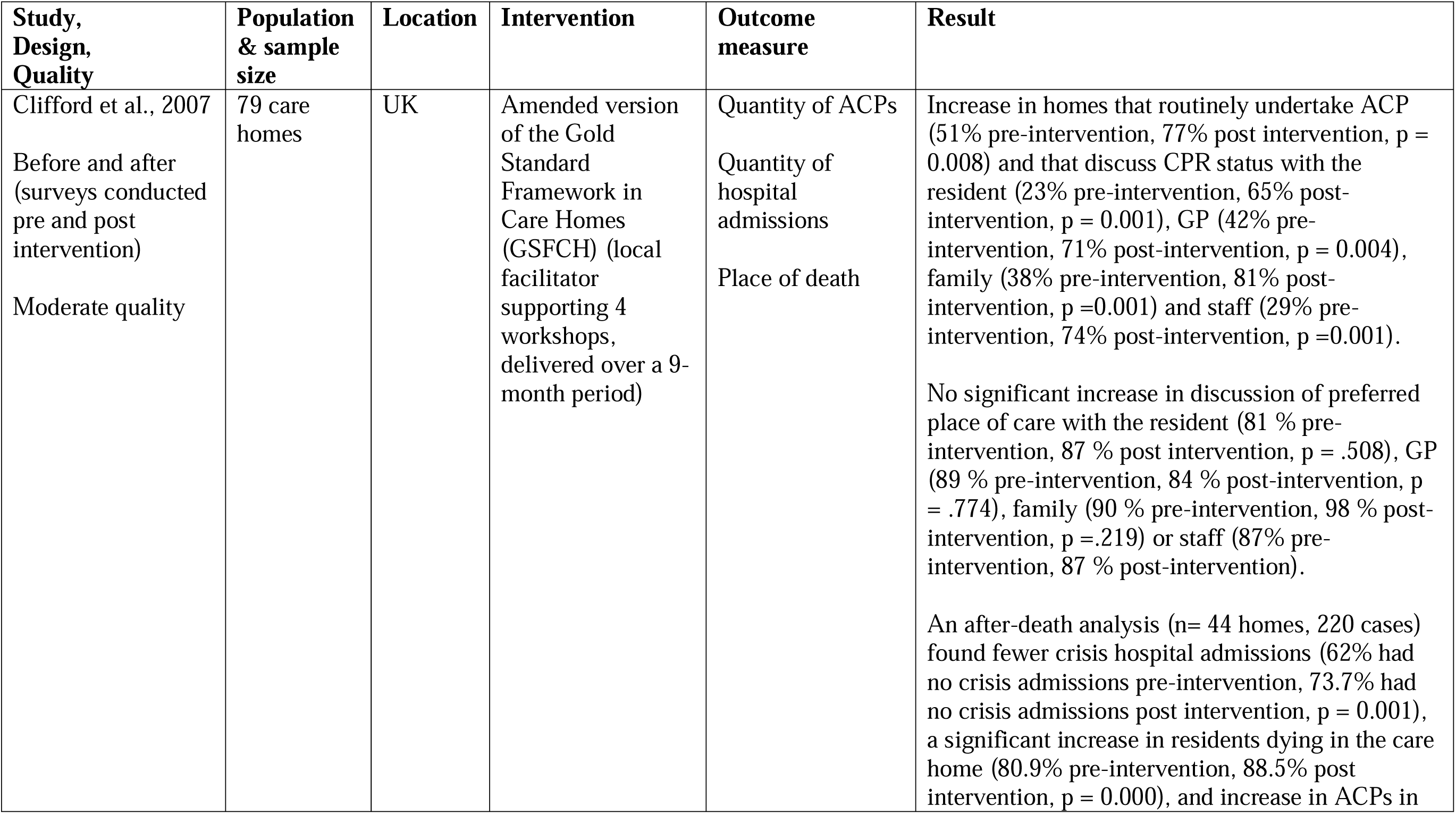
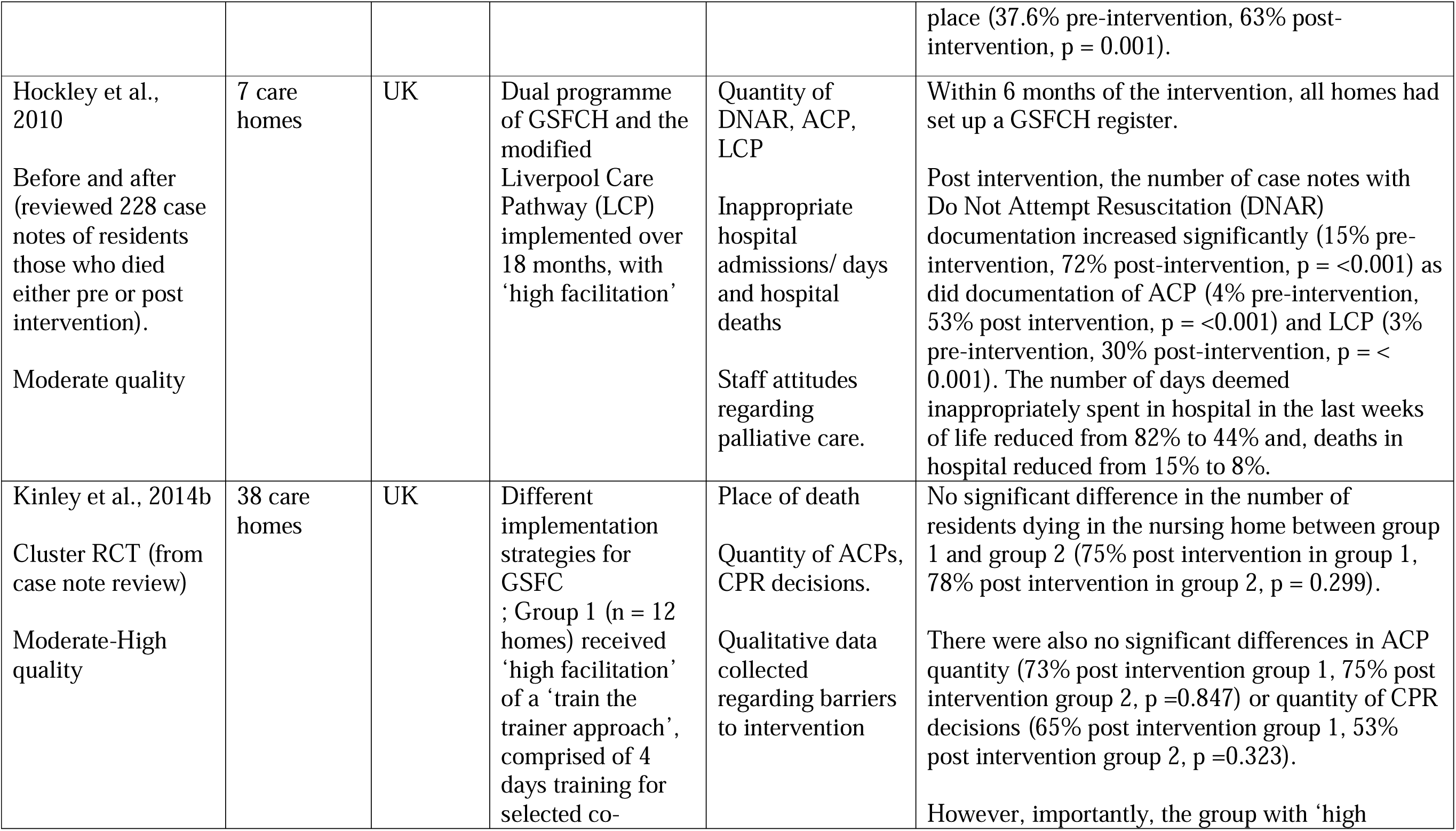
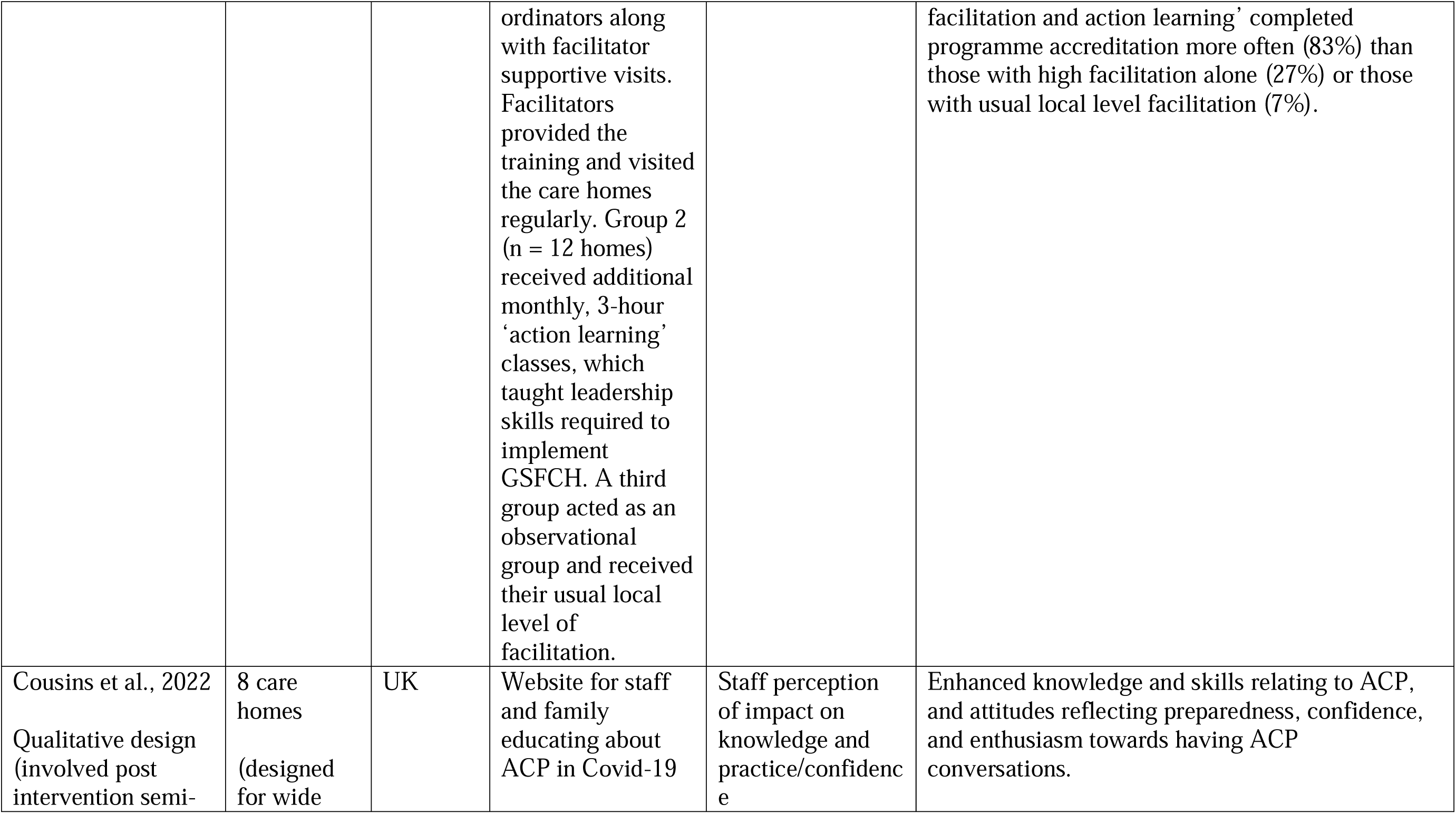
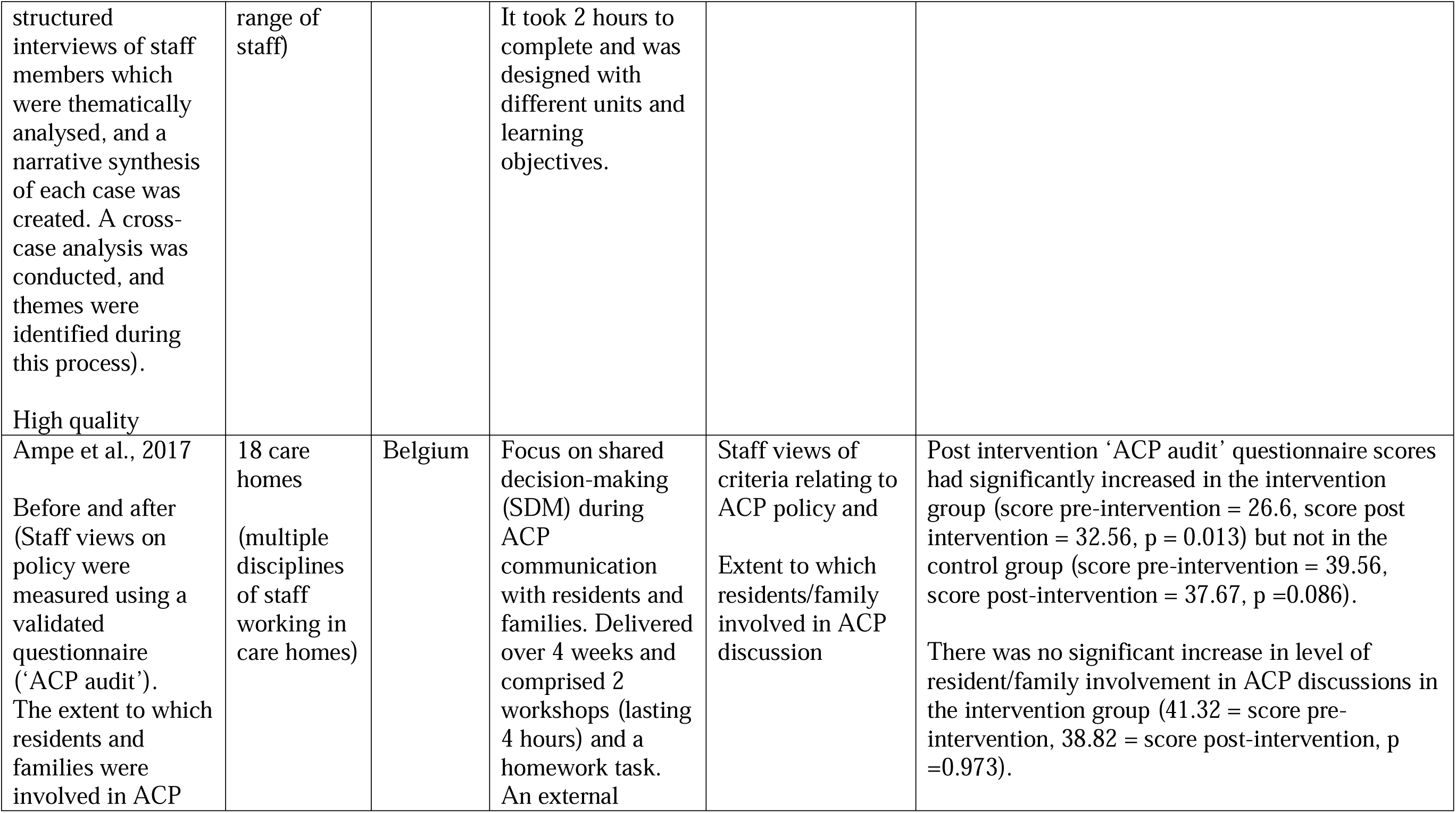
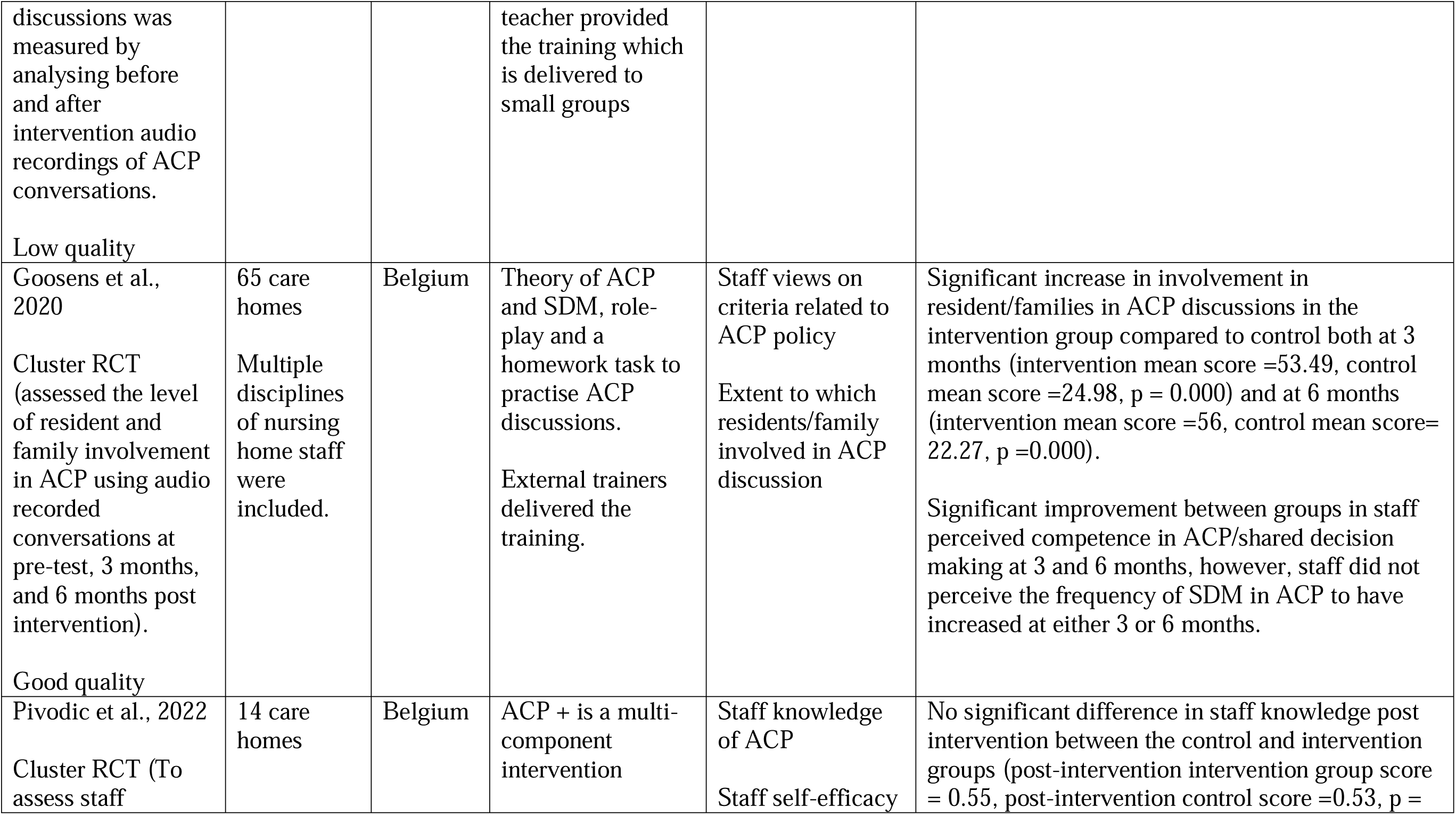
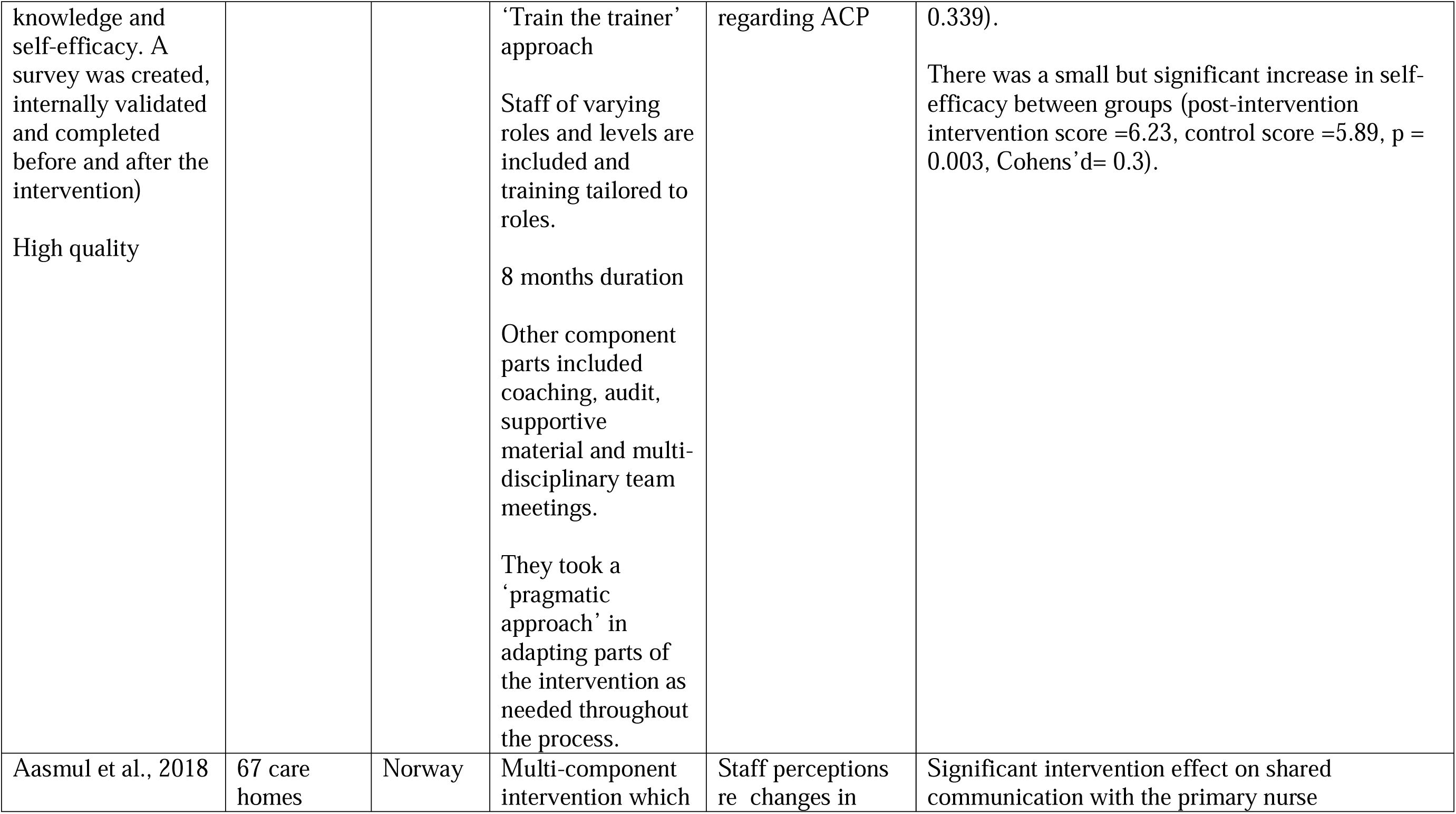
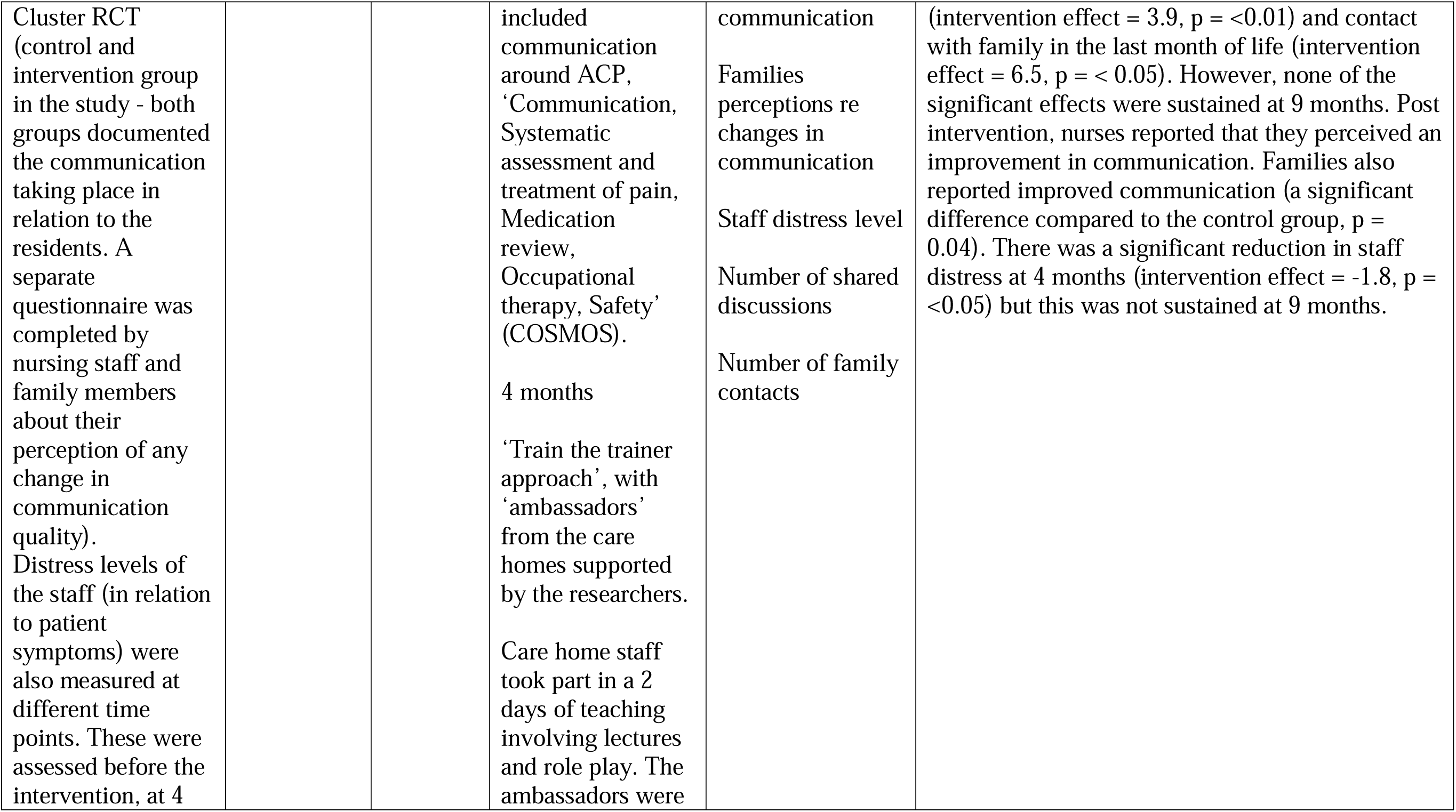
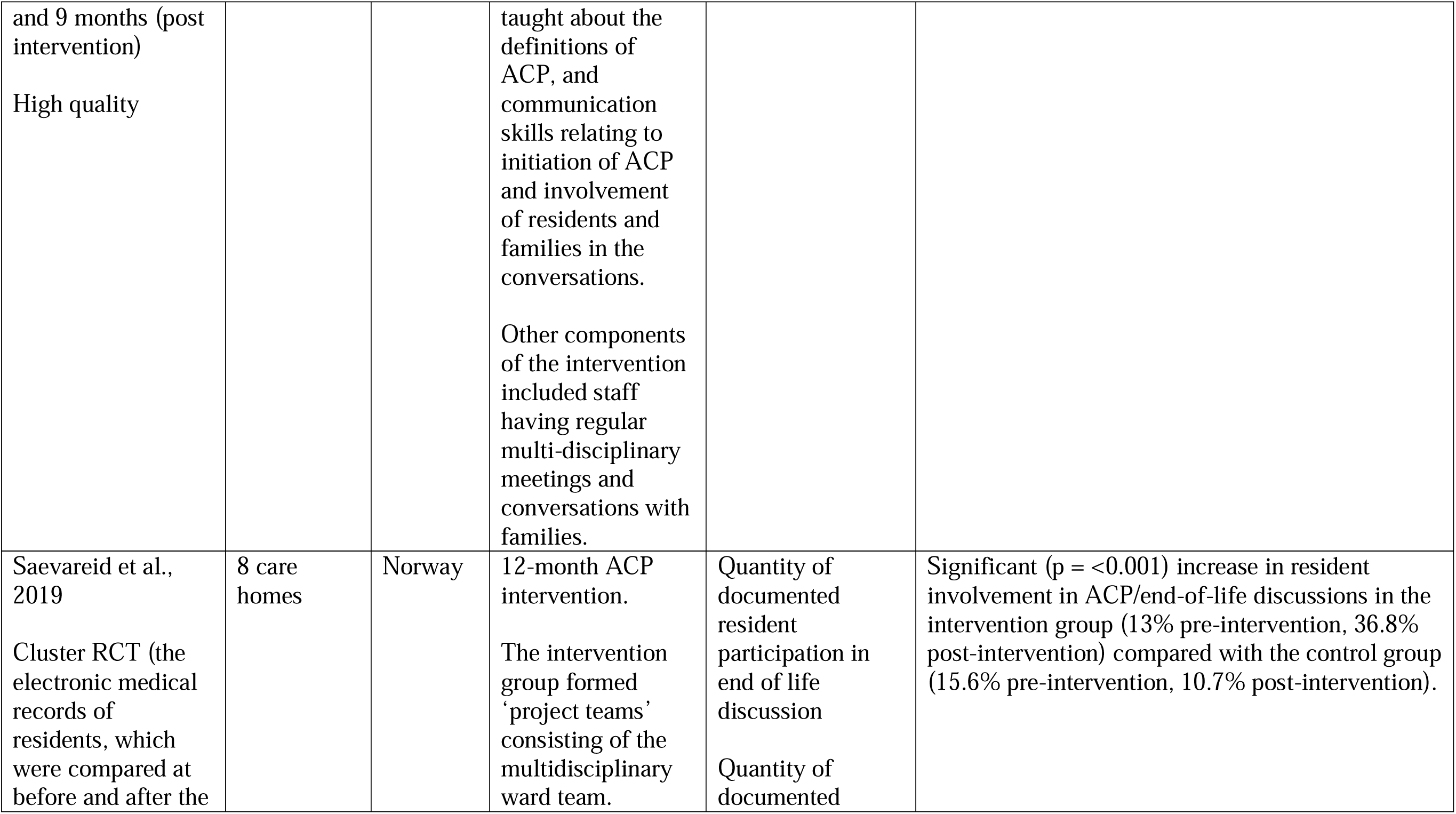
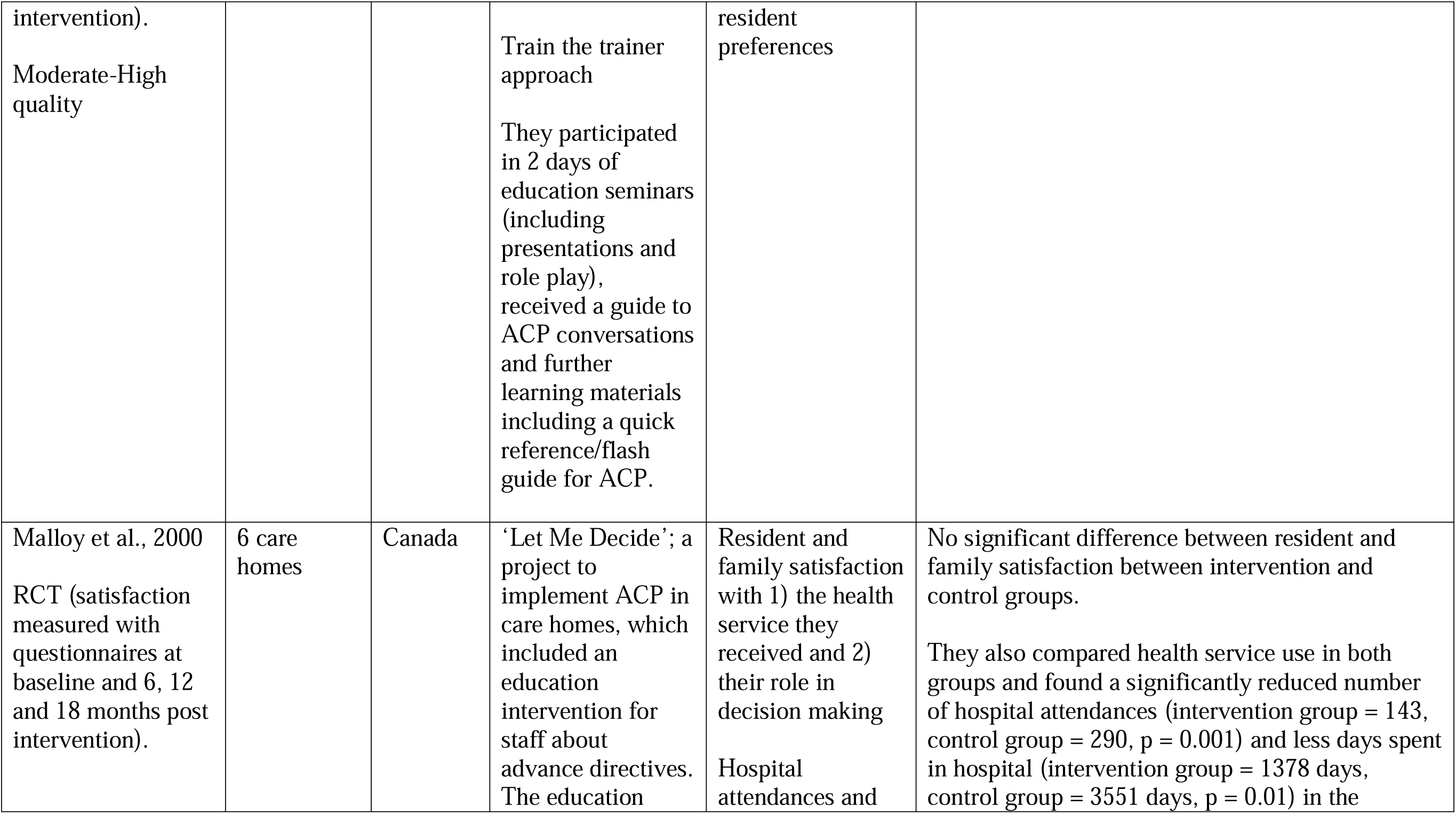
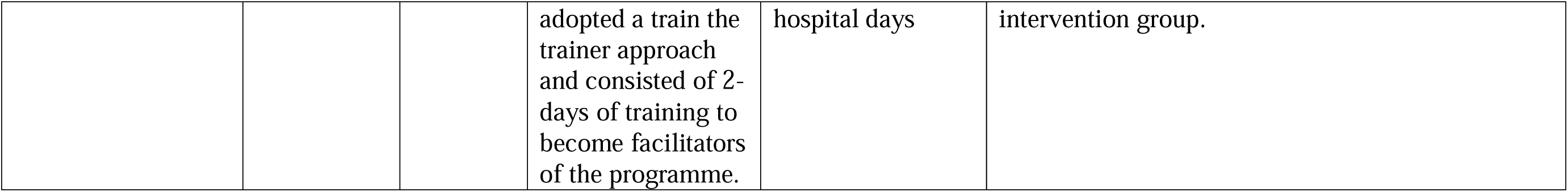
Study Characteristics

**Table 2:** Intervention Design and Outcomes grouped by Country

| Country | Construct of the interventions | Outcomes |
| --- | --- | --- |
| **UK**<br>(n = 4 studies) | 3 studies focus on **The Gold Standards Framework in Care Homes** (GSFCH) - ‘train the trainer’ approach (Hockley et al., 2010; Kinley et al., 2014b) or adoption of local facilitators for training (Clifford et al., 2007).<br>1 study evaluated a **website** intervention for care home staff and families of residents (Cousins et al., 2022) | UK based studies focused mainly on health system/resource –related outcomes: quantity of ACPs (Clifford et al., 2007; Hockley et al., 2010; Kinley et al., 2014b), hospital admissions (Clifford et al., 2007; Hockley et al., 2010), place of death (Clifford et al., 2007; Kinley et al., 2014b). The exception is the recent study by Cousins et al., 2022) which focused on staff and family perceptions of the impact of the intervention. |
| **Belgium**<br>(n = 3 studies) | ‘**We Decide**’ intervention, communication training for shared decision making during ACP (Ampe et al., 2017; Goosens et al., 2020)<br>‘**ACP +**’ is a multi-component intervention, made up of 10 component parts (Pivodic et al., 2022) | Belgian studies focused mainly on staff-related outcomes: staff views of criteria relating to ACP policy (Ampe et al., 2017<sup>34</sup>; Goosens et al., 2020), staff knowledge of ACP and self-efficacy regarding ACP (Pivodic et al., 2022). |
| **Norway**<br>(n = 2) | ‘**Communication, Systematic pain assessment and treatment, Medication review, Organisation of activities and Safety**’ (COSMOS) is a multi-component intervention with a ‘train the trainer’ approach and multiple staff disciplines included in training (Aasmul et al., 2018).<br>Saevareid et al. (2019) evaluated an intervention that involved the whole ward in a nursing home. They adopted a ‘train the trainer’ approach and involved ‘project teams’ consisting of the multidisciplinary team. | Norwegian studies focused on resident/family-related outcomes and staff-related outcomes: staff and family perceptions of changes in communication and staff distress level (Aasmul et al., 2018), quantity of resident participation in end of life discussions (Saevareid et al., 2019). |
| **Canada**<br>(n = 1) | Malloy et al. (2000) evaluated ‘**Let Me Decide**’; a project to implement ACP in care homes, which included an education intervention for staff about advance directives. The education adopted a ‘train the trainer’ approach. | The Canadian study focused on both resident/family-related satisfaction and health system/resource-related hospital admissions (Malloy et al., 2000). |

**Table 3:** Staff Participants in the Primary Studies

| Study | Participants |
| --- | --- |
| Malloy et al.,<br>2000 | • 9 staff (3 for each intervention home)<br>• Nurses |
| Clifford et al.,<br>2007 | • 79 care homes<br>• Care home managers, nurses, carers |
| Hockley et al.,<br>2010 | • Nursing home managers from 7 homes |
| Kinley et al.,<br>2014b | • 24 nursing home managers |
| Ampe et al.,<br>2017 | • 90 staff<br>• All care home staff |
| Aasmul et al.,<br>2018 | • 67 homes<br>• Nurses, doctors, managers |
| Saevareid et al.,<br>2019 | • 8 nursing home wards (4 intervention homes)<br>• Whole wards |
| Goosens et al.,<br>2020 | • 311 staff<br>• All care home staff |
| Pivodic et al.,<br>2022 | • 391 care home staff completed pre and post intervention questionnaires.<br>• Nurses, care assistants, allied health professionals |
| Cousins et al.,<br>2022 | • 35 care staff, 19 family members<br>• Nurses, admin staff, managers, care assistants |

### Risk of Bias Assessment

VBF assessed the quality of each study using the Specialist Unit for Review Evidence Checklist (SURE, 2018) (Table 4). The quality assessments were then checked by AG and any disagreements were resolved by discussion. The SURE (2018) checklists were chosen as they are the updated versions of the tools used in the original review, and they offer a range of tools for risk of bias assessment, which are tailored to the methodology of the primary study, providing separate templates for critical appraisal of experimental studies and qualitative studies (SURE), 2018). In line with the Cochrane Handbook for Systematic reviews of Interventions, (Boutron et al., 2022) the SURE tools allow for systematic consideration of different biases rather than a scoring of the quality.

**Table 4:** Risk of Bias Assessment for Primary Studies

| Study<br>& method | Strength | Reasoning |
| --- | --- | --- |
| Clifford et al. 2007 | Moderate | ✓ Large sample (n = 79 care home)<br>✓ Researchers analysed differences in characteristics between the groups who did and did not complete the surveys.<br>• No control<br>• 54.7% response rate at final audit. |
| Hockley et al. 2010 | Moderate | • Small sample (n = 7 care homes)<br>• No control |
| Kinley et al. 2014b | Moderate-High | ✓ Large sample size (n = 38 care homes).<br>✓ Considered contamination between groups and used the cluster RCT design to overcome this.<br>✓ Groups 1 and 2 were randomised electronically by an external party<br>✓ Data was extracted by two researchers independently, making this process robust<br>• Could not randomise Group 3 who acted as an observational group, which would still be open to confounding |
|  |  | factors. |
| Cousins et al. 2022 | High | ✓ Guided by theoretical propositions.<br>✓ Attrition rate good.<br>✓ Provides rich qualitative data.<br>• Convenience sampling<br>• Small sample (n = 8 care homes) |
| Ampe et al. 2017 | Low | ✓ Moderate sample (n = 18 care homes)<br>• Allocation to intervention and control groups was not random but determined by pre intervention 'ACP audit', in order to identify those care homes with greatest scope for improvement from the intervention. Such a systematic difference between intervention and control groups threatens the internal validity of this study. |
| Goosens et al. 2020 | High | ✓ Groups were randomly allocated before any data was collected and baseline characteristics of participants in the control and intervention groups were similar<br>✓ Large sample (n=65 care homes)<br>• Recruitment method could introduce selection bias as care homes decided which wards to include in the study. |
| Pivodic et al. 2022 | High | ✓ Moderate sample size (n = 14 care homes)<br>✓ Computer generated randomisation of groups to control or intervention.<br>• 50% of respondents completed the post-intervention survey which may produce unreliable results, especially as there is not an analysis of the non-responders |
| Aasmul et al. 2018 | High | ✓ Large sample (n = 67 homes) and covers 3 Norwegian counties.<br>✓ Homes were randomised to control or intervention groups but, this was a constrained process to allow groups to have similar characteristics<br>• The multi-component nature of the intervention makes it difficult to know which elements are effective. |
| Saevareid et al. 2019 | Moderate-High | ✓ Randomised at whole ward rather than individual level -aimed to avoid contamination between groups<br>✓ Pair matched control and intervention groups based on national data<br>✓ Selection bias was minimised by adopting an 'opt out' model. |
|  |  | • Small sample (n = 8 care homes)<br>• First author not blinded to group allocation<br>• Validity of questionnaire used to extract data from case notes not clear |
| Malloy et al. 2000 | Moderate -<br>High | ✓ Pair matched care homes based on characteristics<br>✓ Randomly allocated groups – however, no information given on how the randomisation process worked<br>• Participating care homes were selected to include those with less resident choices documented. This may limit generalisability<br>• Small sample (n = 6 homes) |

### Synthesis Method

Due to the diverse nature of the interventions and outcomes of the studies included in the original review (Gleeson et al., 2021), it was anticipated that the updated results would also not be suitable for meta-analysis, therefore, the plan a priori (Prospero Registration: CRD42022337865), was for the data to be synthesised by narrative synthesis. Further, narrative synthesis allows for studies which are heterogeneous in their method and intervention to be considered in relation to their effectiveness and can produce findings that are accessible for policy and practice (Popay, 2006).

## Results

The update found a further 5 studies that met inclusion criteria, making a total of 10 studies, recruiting staff from 310 care homes. One study (O’Brien et al., 2016) from the original review was excluded, as on further consideration, the intervention focus was not mainly on ACP education. The increase in the number of higher quality studies which met inclusion criteria led to a post hoc decision to exclude new studies of low quality from the review update. As such, all new studies are of moderate-high quality. At the time of the original review, there was less evidence available, and one low quality study was included to provide a more comprehensive review. As suggested by Popay et al. (2006), uncritical inclusion of low quality studies threatens the robustness of the synthesis and studies of equal quality should be given equal weight in the narrative synthesis. To achieve this and ensure internal consistency, the study of low quality by Ampe et al. (2017) is given less weight in the narrative synthesis by acknowledging its methodological flaw and discussing its findings in relation to the same research group’s subsequent, higher quality evaluation of the ACP intervention ‘We Decide’ (Goossens et al., 2020). Figure 1 demonstrates the search strategy. Clear records of decisions for exclusion of full text articles are summarised in Table 5.

**Table 5:** Excluded Studies

|  | Study Reference | Reason for exclusion |
| --- | --- | --- |
| 1. | Flo E, Husebo BS, Bruusgaard P, et al. A review of the implementation and research strategies of advance care planning in nursing homes. *BMC Geriatrics* 2016; 16(24): https://doi.org/10.1186/s12877-016-0179-4 | A review paper |
| 2. | Goossens B, Sevenants A, Declercq A. et al. We DECide optimized' - training nursing home staff in shared decision-making skills for advance care planning conversations in dementia care: protocol of a pretest-posttest cluster randomized trial. *BMC Geriatrics* 2019; 19(1). https://doi.org/10.1186/s12877-019-1044-z | A study protocol |
| 3. | Roh E, Hashimoto T, Samandary S, et al. POLST Training for nurses in nursing homes. *Epidemiology* 2022; 70(suppl 1). https://doi.org/10.1111/jgs.17755 | Abstract only, not able to obtain full text |
| 4. | Katwa A, Jenner C, Ali S. Harrow planning and caring together (PACT) project for care home residents: Development of an inter-professional education programme to improve advance care planning for care home residents. *Age and Ageing* 2018; 47(Suppl 3). https://doi.org/10.1093/ageing/afy126.04 | Abstract only. |
| 5. | Davis J, Morgans A, Dunne M. Supporting adoption of the palliative approach toolkit in residential aged care: an exemplar of organisational facilitation for sustainable quality improvement. | ACP not the overarching focus of education |
|  | Contemporary Nurse: *A Journal for the Australian Nursing Profession* 2019; 55(4/5).<br>https://doi.org/10.1080/10376178.2019.1670708 |  |
| 6. | Hofmann M, Love R, Way D, et al. Improving End-of-Life Care using the Life Sustaining Treatment Decision Initiative in a Veterans Affairs Community Living Centre. *Journal of the American Medical Directors Association* 2020; 21(3).<br>https://doi.org/10.1016/j.jamda.2020.01.062 | Abstract only, not able to obtain full text |
| 7. | Brazil K, Carter G, Cardwell C, et al. *Palliative Medicine* 2017; 32(3).<br>https://doi.org/10.1177/0269216317722413 | Education is not aimed at care home staff |
| 8. | Jurgen. RESPEKT. Study to implement advance care planning (ACP) in the nursing homes (n/h) of a model region by means of qualifying selected n/h staff to facilitate ACP discussions with residents or their proxies. *Cochrane Central Register of Controlled Trials* 2009.<br>https://trialsearch.who.int/Trial2.aspx?TrialID=ISRCTN99887420, Issue 3 | Abstract only, not able to obtain full text |
| 9. | Sawicka Z, Massey K, Carver L. Improving the care of care home residents in wakefield - The wakefield care home vanguard support team. *Age and Ageing* 2017; 46 (suppl 1).<br>https://doi.org/10.1093/ageing/afx055.67 | Not about ACP training |
| 10. | Palmer J, Parker V, Mor V, et al. Barriers and facilitators to | Education is not |
|  | implementing a pragmatic trial to improve advance care planning in the nursing home setting. *BMC health services research* 2019; 19(1). https://doi.org/10.1186/s12913-019-4309-5 | aimed at care home staff |
| 11. | Unroe K, Mitchell, S, Hanson, L, Tu, W, Hickman, S. Pilot of the approaches pragmatic clinical trial-an advance care planning specialist program in nursing homes. *Journal of the American Geriatrics Society* 2019; 67(Suppl 1). https://doi.org/10.1111/jgs.15898 | Abstract only, not able to obtain full text |
| 12. | Kinley J, Hockley J. The sustainability of in-reach end-of-life care programmes into care homes. *Palliative Medicine* 2018; 32(Suppl 1). | Abstract only, not able to obtain full text |
| 13. | Van den Block L, Honinx E, Pivodic L, et al. Evaluation of a palliative care programmed for nursing homes in 7 countries: the PACE cluster-randomised clinical trial. *JAMA International Medicine* 2019; 180(2) 233-242. [doi:10.1001/jamainternmed.2019.5349](https://doi.org/10.1001/jamainternmed.2019.5349) | ACP not the overarching focus of education |
| 14. | Steel A, Hopwood H, Goodwin E, et al. Multidisciplinary residential home intervention to improve outcomes for frail residents. *BMC Health Services Research* 2022; 22(1). https://doi.org/10.1186/s12913-021-07407-y | ACP not the overarching focus of education |
| 15. | Kinley J, Preston N, Froggatt K. Facilitation of an end-of-life care programme into practice within UK nursing care homes: A mixed-methods study. *International Journal of Nursing Studies* 2018; 82. | Outcome is not related to effectiveness |
|  | https://doi.org/10.1016/j.ijnurstu.2018.02.004 |  |
| 16. | Munang L, Rimer J, Ralston K, et al. 164 Standardised Anticipatory Care Planning in Care Homes Reduces Unscheduled Hospital Admissions. *Age and Ageing* 2021; 50 (Suppl 1). https://doi.org/10.1093/ageing/afab030.125 | Abstract only, not able to obtain full text |
| 17. | Di Giulio P, Gonella S. Implementing advance care planning with family caregivers of nursing home residents in Italy. *Palliative Medicine* 2021; 35(Suppl 1). | Abstract only, not able to obtain full text |
| 18. | Park M, Park E, Jo M, et al. Feasibility of an Advance Care Planning Program (ACP) for Korean Community-Dwelling Older Adults and ACP Training of Advance Practice Nurses. *Journal of community health nursing* 2021; 38(3).<br>https://doi.org/10.1080/07370016.2021.1932963 | Not about care homes |
| 19. | Gleeson A, Noble S, Mann M. Advance care planning for home health staff: a systematic review. *BMJ supportive & palliative care* 2021; 11(2). | Review paper |
| 20. | Rainsford S, Liu W, Johnston N, et al. The impact of introducing Palliative Care Needs Rounds into rural residential aged care: A quasi-experimental study. *The Australian journal of rural health* 2020; 28(5). https://doi.org/10.1111/ajr.12654 | ACP not the overarching focus of education |
| 21. | Phua G, Li G, Toh H, et al. Educational needs of nursing home staff: does a needs-based palliative care course make a difference? *BMJ supportive & palliative care* 2020; doi: 10.1136/bmjspcare-2020-002690 | ACP not the overarching focus of education |
| 22. | Sun A, Crick M, Orosz Z, et al. An Evaluation of the | ACP not the |
|  | Communication at End-of-Life Education Program for Personal Support Workers in Long-Term Care. *Journal of palliative medicine* 2022; 25(1). https://doi.org/10.1089/jpm.2021.0054 | overarching focus of education |
| 23. | Coalition Corner: Health Care Decisions Coalition. *New Hampshire Nursing News* 2021; 45(4) | Abstract only, not able to obtain full text |
| 24. | Rockingham Heroes. *New Hampshire Nursing News*. 2021. | Abstract only, not able to obtain full text |
| 25. | Livingston G, Lewis-Holmes E, Pitfield C, et al. Improving the end of life for people with dementia living in a care home: an intervention study. *International Psychogeriatrics* 2013; 25(11). doi:10.1017/S1041610213001221 | Low Quality |
| 26. | Fernandez-Rodriguez A, Molina-Mula J, Sarabia-Cobo C. Education intervention: improving the knowledge and attitudes of health professionals on living wills. *Nurse Education Today* 2021; DOI: [10.1016/j.nedt.2021.105016](https://doi.org/10.1016/j.nedt.2021.105016) | Low Quality |
| 27. | Wils M, Verbakel J, Lisaerde, J. Improving advance care planning in patients with dementia: the effect of training nurses to engage in ACP-related conversations. *Journal of Clinical Gerontology & Geriatrics* 2017; 8(1):17-20 | Low Quality |
| 28. | Katwa A., Jenner C, MacDonald K, et al. Improving advance care planning for care home residents with dementia: Evaluation of simulation training for care home workers. *Dementia* 2020; 19(3). | Low Quality |
|  | doi: 10.1177/1471301218788137. |  |
| 29. | Kezirian A, McGregor M, Stead U, et al. Advance Care Planning in the Nursing Home Setting: A Practice Improvement Evaluation. *Journal of Social Work in End of Life & Palliative Care* 2018; 14(4). doi: 10.1080/15524256.2018.1547673. | Low Quality |
| 30. | O'Brien M, Kirton J, Knighting K, et al. Improving end of life care in care homes; an evaluation of the six steps to success programme. *BMC Palliative Care* 2016; 15(1):53. | ACP not the overarching focus of education |

## Narrative Synthesis

The framework for narrative synthesis of systematic reviews by Popay et al. (2006) is adopted here, as it gives structure to syntheses which focus on the effectiveness of interventions. The characteristics of this framework are: 1) consider the interventions “theory of change”, 2) “develop a preliminary synthesis”, 3) “explore relationships within and between studies” and 4) “assess the robustness of the synthesis”. The robustness of the synthesis is discussed in the broader context of review strengths and limitations in the discussion section.

### 1. Consider the intervention’s ‘theory of change’

The “theory of change” can be considered as an explanation of how an intervention has impact (Popay, 2006). This is explicitly explored by Pivodic et al. (2022) who conducted a literature review and stakeholder engagement to inform their theory that care home staff must have adequate self-efficacy (confidence in their ACP-related abilities) and knowledge regarding ACP, in order to make improvements in the ACP process in care homes. This theory also underpins the studies by Saevaried et al. (2019) and Cousins et al. (2022), who believe that increased knowledge/competence in relation to ACP would improve its uptake. Aasmul et al. (2018) consider that if care home staff don’t feel competent in managing complex care home residents, they may experience subsequent distress. They hypothesise that ACP education will therefore, relieve staff distress. They base this theory on results of a study demonstrating that care home nursing aids experienced reduced caregiver burden, compared to a control group, when they were trained in communication skills (Sprangers et al., 2015). Goosens et al. (2020) argue that communication skills without self-efficacy are not likely to be sufficient to influence positive change. This is based on evidence that lack of self-efficacy regarding ACP presents a barrier to ACP in care homes (Harrison Dening et al., 2019).

### 2. Develop a Preliminary Synthesis

Initially, a descriptive paragraph was written, summarising each of the studies. Studies were then grouped by country to allow for consideration of the applicability of the results to different countries, in recognition of the associated different health care systems, resources and legal frameworks around death and dying. These confounding factors result it not being possible to assume comparable effectiveness of the interventions across countries (Lavis et al., 2009). Further, organising by intervention/method/outcome measure/results is not practical here, due to the heterogeneity of these elements. The summarised data were then tabulated (Table 1) to visualise the study characteristics.

### 3. Explore relationships within and between studies

Major sources of heterogeneity between studies should be scrutinised as to whether they act as mediating factors, understood as those which influence the results of primary studies (Popay, 2006). A systematic approach to identifying potential relationships within and between primary studies is ‘ideas webbing’, which allows visualisation of relationships in the data (Popay, 2006). This approach was adopted here as it grouped studies together, allowing identification of the following potential mediating factors: intervention, population and outcomes. Visualisation of the relationships identified between the mediating factors during the ideas webbing process is demonstrated in Supplementary File 2. The potential impact of intervention, population and outcome heterogeneity on the results will be analysed as the major foci of the narrative synthesis.

### Variability in Interventions

#### ACP Interventions are Complex Interventions

Recent Medical Research Council guidance (Skivington et al., 2021) suggests that interventions can be considered as “complex interventions” when they have multiple component parts (e.g. Aasmul et al., 2018; Pivodic et al., 2022), target multiple different levels of participant (e.g. Ampe et al., 2017, Aasmul et al., 2018, Saevareid et al., 2019, Goosens et al, 2020, Cousins et al., 2022 and Pivodick et al., 2022) and require a high level of skill for delivery and uptake. Further, flexible interventions (e.g. Pivodic et al., 2022) can be considered as complex, as decisions need to be made about which components of the intervention are flexible and which are fixed. These elements which add complexity and are present in many of the ACP education interventions.

#### Multi-Component versus Single-Component Interventions

Despite broadly similar theories of change, the studies have diverse designs. Some interventions are multi component (e.g. Aasmul et al., 2018 and Pivodic et al., 2022), while others are single component. The impact of this on the results is considered by Pivodic et al., 2022, who suggest that outcomes may be more difficult to achieve and complex to interpret when an intervention has multiple component parts. This may explain Pivodic et al.’s relatively neutral results, however, Aasmul et al., 2018 found significant intervention effects, despite their multi-component design. It may be that the difference here is related to the different outcome measures adopted by the studies; Aasmul et al., 2018 adopted staff-related, resident/family related and health resource related outcomes whereas Pivodic et al., 2022 considered only staff related outcomes. They also had different follow up periods which may be relevant as Aasmul et al.’s intervention effect was not sustained at 9 month follow up.

#### Train the trainer

All countries had studies which adopted a ‘train the trainer’ approach. It may be that this approach allows for improved sustainability of the intervention impact, (Health Education England., 2017) as the care home staff continue to cascade knowledge, however, follow up of these interventions is needed to better understand this. Further, this approach assumes that staff feel competent and have the time and resources to share their learning with others (Health Education England., 2017). This adds complexity to the intervention as the staff must have the expertise to both learn and teach new skills (Skivington et al., 2021). It may be useful to explore staff views on this element of the interventions with future qualitative research.

#### Flexible Interventions

Pivodic et al. (2022) allowed their intervention to be tailored to the care home as it was rolled out. Changes were made to the number of meetings held, roles of staff, timings for education sessions etc. Importantly, they agreed on a list of flexible and fixed elements of the intervention at the beginning of the process, which may be important for generalisability to other settings.

### Variability in Populations

The care home staff participants in the studies have extended in breath over time, with the earlier education interventions targeting only nursing home managers (Hockley et al., 2010, Kinley et al., 2014b) or nurses (Malloy et al., 2000). Many studies mention the importance of involving nursing home management in the intervention (Kinley et al., 2014b, Hockley et al., 2010, Ampe et al., 2017, Aasmul et al., 2018, Goosens et al., 2020) in order to ensure time is allocated for the training, and the culture of the home is receptive to change. The more recent studies target the broader multi-disciplinary care home team, involving a range of staff disciplines (Ampe et al., 2017, Aasmul et al., 2018, Saevareid et al., 2019, Goosens et al., 2020, Cousins et al., 2022, Pivodic et al., 2022). This reflects the need for all resident and family-facing staff to have familiarity with the ACP process in order to support its implementation (Goossens et al., 2020). Saevareid et al. (2019) trained whole wards within the care homes which they argue improves internal validity by reducing contamination of control wards and reduces participant selection bias. Further, Cousins et al. (2022) included family members alongside staff as participants in their educational intervention. The staff participants in the primary studies are summarised in Table 3.

Pivodic et al. (2022) delivered tailored training to different staff disciplines and allocated them specific roles in the role out of their education intervention. This may more closely meet the training needs of the staff members. In terms of analysing the results, Pivodic et al. mention that their results are grouped, rather than considered separately for different staff disciplines, which may have influenced their outcomes. It may be more relevant to consider results at staff discipline level (in relation to their specific role) rather than to group results, as this would better reflect the knowledge or self-efficacy required by the different staff disciplines.

### Variability in Outcomes

This update found that all of the primary studies adopt multiple outcome indicators. These are diverse, however, can be categorised into constructs. The outcome categories that emerged during the ideas webbing process include resident/family-related, staff-related and health system/resource related construct outcomes. Bearing in mind all studies have more than one outcome measure, it was possible to broadly group outcome measure by country (Table 2).

## Discussion

To the best of our knowledge, this is the most up-to-date systematic review of ACP education interventions for care home staff. ACP education interventions can be considered as complex interventions as they can be multi-component, flexible and target multiple levels of participants. These factors which add complexity, reflect the heterogeneity in the studies intervention designs and may act as mediating factors that impact on the results. Further heterogeneity between studies relates to the diverse outcome measures adopted.

What constitutes positive impact of an ACP education intervention is muddied, partly due to the voluntary nature of ACP (Pivodic et al., 2022). It is not compulsory to complete an ACP, and as such, for a minority of people, not completing an ACP is their choice, and arguably a measure of success. It is therefore too simplistic to assume that a higher quantity of ACPs is equal to positive impact. Similarly, documentation of ACP is not equivocal to evidence of its being used effectively in decision making (Flo et al., 2016) and may be too simplistic an outcome.

Another common outcome measure adopted in the primary studies is “number of residents who died in the care home” (e.g. Clifford et al., 2007, Kinley et al., 2014b). It is not clear if this is a positive or negative outcome, unless this is measured in the context of the residents preferred place of death. Clifford et al. (2007) found that the intervention was associated with more residents dying in the care home, but not with increased discussion of preferred place of death. Arguably, preferred place of death is more important to understand in order to provide person centred care.

An international Delphi study (Rietjens et al., 2017) provides agreement and consensus on which outcome measure constructs should be adopted when evaluating ACP. Their rigorous technique, which included the opinions of over 100 experts, involved patient representatives, and a meta- review of the literature, resulted in 14 constructs being recommended. Of these, “self-efficacy to engage in ACP” (staff, individuals and family) and “use of health care” both received very strong consensus. Our review adds the latest evidence of which outcome measures are in use in relation to ACP education interventions in care homes. The question now arises as to how to move forward with the recommendations from the international Delphi study and the most recent evidence of the outcome measures being adopted.

The Medical Research Council (Skivington et al., 2021) suggest that when researching complex interventions, considering the outcome alone is not sufficient, and that understanding impact mechanism (or ‘theory of change’), and the interaction of the intervention with its context are other important factors to consider. Another approach may therefore be to revisit the ‘theory of change’ that underpins the intervention impact. From the included primary studies, a possible, common theory of change is that both subjective (self-efficacy) and objective (knowledge) changes may be needed in order to achieve positive behaviour change relating to ACP practice. Self-efficacy is therefore both recommended by the Delphi study (Rietjens et al., 2017) and the above theory of change as being an important construct to consider when measuring ACP interventions. The studies which consider staff self-efficacy-related outcome measures in this review include Goosens et al., 2020 (demonstrated increased perceived competence in shared decision making in the intervention group), Cousins et al., 2022 (demonstrated increased staff and family confidence for ACP), Pivodic et al., 2022 (demonstrated increased staff self-efficacy regarding ACP). Of note the results of these studies in relation to self-efficacy all indicate positive change in relation to the interventions. However, Goosens et al. (2020) found that their intervention significantly increased staff perceived competence in shared decision making, but did not influence staff perceived use of shared decision making and Pivodic et al., 2022 found that their intervention improved staff self-efficacy but not engagement in ACP. These results suggest that self-efficacy alone may not be sufficient for positive behaviour change.

### Strengths and Limitations

The review is strengthened by its detailed protocol, strict adherence to the PRISMA 2020 guidance and the framework adopted for narrative synthesis. A search of multiple databases, journals and websites was conducted in attempt to access all relevant literature. Two authors independently screened studies for inclusion. Systematic assessment of the methodological quality of the primary studies using the Specialist Unit for Review Evidence Guidelines (2018) ensured robust quality assessment of the studies. Equal weight is given to studies of similar quality (Popay, 2006), with Ampe et al. (2017) considered in relation to its methodological flaw and alongside this research teams’ subsequent paper (Goosens et al., 2020).

A limitation of this review is that it was not possible to conduct a meta-analysis of the data. However, Popay et al.’s framework for narrative synthesis was adopted in order to have a rigorous and structured process to synthesis. Further, the different countries and contexts in which the interventions took place limit generalisability out with their countries of origin. However, the global search for studies aimed to enhance generalisability of the results by having a geographical spread of primary studies. The review is limited as it includes only studies in English language. Finally, despite the relatively short time period between the original search and review update (4 years), the review update almost doubled the number of included studies, and as such provides novel findings.

### Implications Future Research and Clinical Practice

The following points are relevant to future research involving complex ACP education interventions and are in keeping with the Medical Research Council’s framework for developing and evaluating complex interventions (Skivington et al., 2021).

#### Intervention Design

- When targeting multiple staff disciplines, consider the impact of grouping results and possible benefits of measuring outcomes in a way that reflects the education needs of staff disciplines.
- Involvement of care home management seems key for interventions to be implemented successfully.
- If adopting a flexible intervention design which is adapted during the intervention, consider listing flexible and fixed elements of the intervention in the study protocol. This would aid subsequent generalisability of the results to other groups.
- There is a lack of evidence relating to the longer term impact of ACP education interventions. A considered follow up would give insight into the sustainability of the interventions and guide ongoing education.

#### Clinical Practice

- If adopting a ‘train the trainer’ approach, consider the skills required for the staff to learn and then subsequently teach the material. Gain insight into how care home staff feel about cascading knowledge.
- Consider a pooled, co-ordinated approach to education across care homes, which may potentially limit the impact of training being diluted by staff attrition and sustain the ‘train the trainer’ approach.
- When ACP training interventions are introduced, ensure that service managers use suitable outcome measures.
- Aim to build care home staff knowledge and self-efficacy regarding ACP.

#### Choice of Outcome measures

- Consider updating the Delphi process (Rietjens et al., 2017) in light of the systematically collected evidence provided here, with a view to agreeing outcomes that are specific to ACP education interventions for care home staff.
- Intervention studies could consider their theory of change to aid decisions about which outcome measures to adopt.
- It may be that that both subjective (self-efficacy) and objective (knowledge) changes are needed in order to achieve positive behaviour change relating to ACP practice.

## Conclusion

The current review update almost doubled the number of included studies in a relatively short time frame, demonstrating that this is a rapidly evolving field of research. ACP education interventions are heterogeneous and complex in their multi-component design, flexibility, different target populations, and outcomes. The Medical Research Council recommends that such complexity may require consideration of the interventions ‘theory of change’. Outcome measures are still diverse, and commonly employed outcome measures, such as quantity of ACP and number of nursing home deaths, may be too simplistic and not reflect the genuine wishes of the resident. Considering the interventions ‘theory of change’, and the international Delphi consensus, it may be that both staff self-efficacy and knowledge in relation to ACP are important outcomes to consider. Future research could consider updating the Delphi process in light of the systematically collected evidence provided here, with a view to agreeing outcomes that are specific to ACP education interventions for care home staff.

## Data Availability

Not relevant as this is a systematic review

## Acknowledgements

I would like to thank Marshall Dozier and Dr Ruth McQuillan (Usher Institute, Uncover Team, University of Edinburgh) who provided technical guidance and support throughout the systematic review process.

