## Supplementary File 1 for "The Effectiveness of Advance Care Planning Training for Care Home Staff: a Systematic Review"

Search Name:	COCHRANE COMBINED TUESDAY 14/6

Date Run:	14/06/2022 21:55:26

Comment:	Limited from March 2018-June 2022

Added MeSH nursing assistant

ID	Search	Hits

#1	MeSH descriptor: [Advance Directives] explode all trees	126

#2	MeSH descriptor: [Advance Care Planning] explode all trees	302

#3	"anticipatory care plan*" or "advanced care plan*" or "advance directive*" or "living will" or "right to die"	1146

#4	#1 or #2 or #3	1336

#5	MeSH descriptor: [Homes for the Aged] explode all trees	663

#6	MeSH descriptor: [Nursing Homes] explode all trees	1543

#7	"nursing home*" or "residential home for the elderly" or "long-term care facilities" or "residential care" or "care home*"	6002

#8	#5 or #6 or #7	6447

#9	MeSH descriptor: [Education] explode all trees	35530

#10	train* or education or learn*	215548

#11	#9 or #10	223291

#12	nurse* or "health care assistant" or "allied health" or doctor* or physician* or "nursing staff" or "nursing personnel" or therapist* or physiotherapist* or "health care professional*"	114533

#13	MeSH descriptor: [Nurses] explode all trees	1319

#14	MeSH descriptor: [Nursing Assistants] explode all trees	74

#15	MeSH descriptor: [Physicians] explode all trees	2401

#16	#12 or #13 or # 14 or #15	354797

#17	#4 and #8 and #11 and #16 with Cochrane Library publication date Between Mar 2018 and Jun 2022	70

ERIC. 14/6/22

Didn’t make changes to original search other than to add limitation of date added to Eric.

No results.

| S9 | S3 AND S4 AND S7 AND S8 AND (da: 2018 OR 2019 OR 2020 OR 2021 OR 2022) | **Expanders** - Apply equivalent subjects  **Search modes** - Boolean/Phrase | [**View Results**](javascript:__doPostBack('ctl00$ctl00$MainContentArea$MainContentArea$historyControl$HistoryRepeater$ctl00$linkResults','')) (0)  [**View Details**](javascript:showShDetails(%22ctl00_ctl00_MainContentArea_MainContentArea_historyControl_ctrlPopup%22,%20%22S9%22,%20true);)  [**Edit**](https://web.s.ebscohost.com/Legacy/Views/UserControls/Ehost/) |  |
| --- | --- | --- | --- | --- |
| 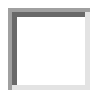 | S8 | S6 OR S7 | **Expanders** - Apply equivalent subjects  **Search modes** - Boolean/Phrase | [**Rerun**](javascript:__doPostBack('ctl00$ctl00$MainContentArea$MainContentArea$historyControl$HistoryRepeater$ctl01$linkResults',''))  [**View Details**](javascript:showShDetails(%22ctl00_ctl00_MainContentArea_MainContentArea_historyControl_ctrlPopup%22,%20%22S8%22,%20true);)  [**Edit**](https://web.s.ebscohost.com/Legacy/Views/UserControls/Ehost/) |
| 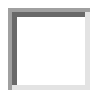 | S7 | TX nurse* or "health care assistant" or doctor* | **Expanders** - Apply equivalent subjects  **Search modes** - Boolean/Phrase | [**Rerun**](javascript:__doPostBack('ctl00$ctl00$MainContentArea$MainContentArea$historyControl$HistoryRepeater$ctl02$linkResults',''))  [**View Details**](javascript:showShDetails(%22ctl00_ctl00_MainContentArea_MainContentArea_historyControl_ctrlPopup%22,%20%22S7%22,%20true);)  [**Edit**](https://web.s.ebscohost.com/Legacy/Views/UserControls/Ehost/) |
| 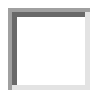 | S6 | ( "allied health" or doctor* or nurse or nurses or physician* or nursing staff or nursing personnel or therapist* or physiotherapist* or" health care professional*" | **Expanders** - Apply equivalent subjects  **Search modes** - Boolean/Phrase | [**Rerun**](javascript:__doPostBack('ctl00$ctl00$MainContentArea$MainContentArea$historyControl$HistoryRepeater$ctl03$linkResults',''))  [**View Details**](javascript:showShDetails(%22ctl00_ctl00_MainContentArea_MainContentArea_historyControl_ctrlPopup%22,%20%22S6%22,%20true);)  [**Edit**](https://web.s.ebscohost.com/Legacy/Views/UserControls/Ehost/) |
| 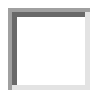 | S5 | TX "nursing home*" OR AB "residential home for the elderly" OR AB "long-term care facilities" OR AB "residential care" OR TX "care home*" | **Expanders** - Apply equivalent subjects  **Search modes** - Boolean/Phrase | [**Rerun**](javascript:__doPostBack('ctl00$ctl00$MainContentArea$MainContentArea$historyControl$HistoryRepeater$ctl04$linkResults',''))  [**View Details**](javascript:showShDetails(%22ctl00_ctl00_MainContentArea_MainContentArea_historyControl_ctrlPopup%22,%20%22S5%22,%20true);)  [**Edit**](https://web.s.ebscohost.com/Legacy/Views/UserControls/Ehost/) |
| 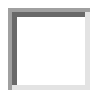 | S4 | TX train* OR TX education OR TX learn* | **Expanders** - Apply equivalent subjects  **Search modes** - Boolean/Phrase | [**Rerun**](javascript:__doPostBack('ctl00$ctl00$MainContentArea$MainContentArea$historyControl$HistoryRepeater$ctl05$linkResults',''))  [**View Details**](javascript:showShDetails(%22ctl00_ctl00_MainContentArea_MainContentArea_historyControl_ctrlPopup%22,%20%22S4%22,%20true);)  [**Edit**](https://web.s.ebscohost.com/Legacy/Views/UserControls/Ehost/) |
| 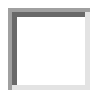 | S3 | S1 OR S2 | **Expanders** - Apply equivalent subjects  **Search modes** - Boolean/Phrase | [**Rerun**](javascript:__doPostBack('ctl00$ctl00$MainContentArea$MainContentArea$historyControl$HistoryRepeater$ctl06$linkResults',''))  [**View Details**](javascript:showShDetails(%22ctl00_ctl00_MainContentArea_MainContentArea_historyControl_ctrlPopup%22,%20%22S3%22,%20true);)  [**Edit**](https://web.s.ebscohost.com/Legacy/Views/UserControls/Ehost/) |
| 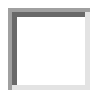 | S2 | AB ( "end of life" or EOL ) AND AB ( care or discuss* or decision* or plan or plans or planning or preference* ) | **Expanders** - Apply equivalent subjects  **Search modes** - Boolean/Phrase | [**Rerun**](javascript:__doPostBack('ctl00$ctl00$MainContentArea$MainContentArea$historyControl$HistoryRepeater$ctl07$linkResults',''))  [**View Details**](javascript:showShDetails(%22ctl00_ctl00_MainContentArea_MainContentArea_historyControl_ctrlPopup%22,%20%22S2%22,%20true);)  [**Edit**](https://web.s.ebscohost.com/Legacy/Views/UserControls/Ehost/) |
| 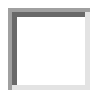 | S1 | TX "anticipatory care plan*" OR TX "advanced care plan*" OR TX "advance directive*" OR AB "living will" OR AB right to die | **Expanders** - Apply equivalent subjects  **Search modes** - Boolean/Phrase |  |

Embase <1980 to 2022 Week 23>

Subject headings added to include: Nursing assistant/ or nursing aide/ and nursing home or nursing homes/

1	advance care planning/	4656

2	(advance adj (care or health or medical) adj plan*).mp.	7602

3	((advance or anticipatory) adj (plan or plans or planning)).tw.	553

4	(advance adj (directive* or decision*)).tw.	5588

5	anticipatory care plan*.tw.	94

6	living will/	9505

7	(living will or right to die).tw.	1914

8	((end of life or EOL) adj5 (care or discuss* or decision* or plan or plans or planning or preference*)).tw.	25952

9	(Advance Care Plan* or ACP).tw.	16460

10	or/1-9	49684

11	health care personnel/	202582

12	physicians/	201435

13	nurse/	136387

14	nursing assistant/ or nursing aide/	4699

15	(nurse* or health care assistant or doctor*).tw.	519476

16	(allied health or doctor* or nurse or nurses or physician* or nursing staff or nursing personnel or therapist* or physiotherapist* or health care professional*).tw.	1138163

17	((health or medical) adj (staff or worker* or professional or personnel or provider or practitioner)).tw.	81047

18	or/11-17	1430984

19	Education/	432575

20	(teach* or educat* or learn* or instruct* or train*).tw.	2217572

21	Learning/	219714

22	or/19-21	2381103

23	nursing home/ or nursing homes/	54743

24	residential home/	7196

25	assisted living facility/	2935

26	long term care/	137616

27	nursing home*1.tw.	41587

28	care home*1.tw.	6141

29	((elderly or old age) adj2 home*1).tw.	2875

30	assisted living facilit*.tw.	1209

31	((nursing or residential) adj (home*1 or facilit*)).tw.	49984

32	(home*1 for the aged or home*1 for the elderly or home*1 for older adult*).tw.	3605

33	residential aged care.tw.	1616

34	("frail elderly" adj2 (facilit* or home or homes)).tw.	87

35	(residential adj (care or facilit* or setting*)).tw.	6996

36	or/23-35	218186

37	10 and 18 and 22 and 36	659

38	limit 37 to (english language and dc=20180301-20220606)	219

39	limit 38 to dc=20180301-20220614	219

Ovid MEDLINE(R) <1946 to June Week 1 2022>

1	Advance Care Planning/	3849

2	(advance adj (care or health or medical) adj plan*).mp.	5144

3	((advance or anticipatory) adj (plan or plans or planning)).tw.	363

4	Advance Care Plan*.tw.	3407

5	(advance adj (directive* or decision*)).tw.	3829

6	anticipatory care plan*.tw.	29

7	Advance Directives/	6427

8	living will*.tw.	1243

9	Right to Die/	4958

10	right to die.tw.	918

11	ACP.tw.	8742

12	((end of life or EOL) adj5 (care or discuss* or decision* or plan or plans or planning or preference*)).tw.	15800

13	or/1-12	36571

14	Health Personnel/	57728

15	Physicians/	98038

16	Nurses/	43728

17	Nursing assistants/ or Nurses Aides/	4353

18	(nurse* or health care assistant or doctor*).tw.	383513

19	(allied health or doctor* or nurse or nurses or physician* or nursing staff or nursing personnel or therapist* or physiotherapist* or health care professional*).tw.	775618

20	((health or medical) adj (staff or worker* or professional or personnel or provider or practitioner)).tw.	56089

21	or/14-20	916636

22	Education/	21496

23	(teach* or educat* or learn* or instruct* or train*).tw.	1474893

24	Learning/	75336

25	or/22-24	1497340

26	exp nursing homes/	43015

27	Residential Facilities/	5703

28	Assisted Living Facilities/	1552

29	Long-Term Care/	27709

30	nursing home*1.tw.	30694

31	care home*1.tw.	4014

32	((elderly or old age) adj2 home*1).tw.	2076

33	assisted living facilit*.tw.	752

34	((nursing or residential) adj (home*1 or facilit*)).tw.	35711

35	(home*1 for the aged or home*1 for the elderly or home*1 for older adult*).tw.	3001

36	residential aged care.tw.	1256

37	("frail elderly" adj2 (facilit* or home or homes)).tw.	63

38	(residential adj (care or facilit* or setting*)).tw.	5147

39	or/26-38	89133

40	13 and 21 and 25 and 39	315

41	limit 40 to dt=20180301-20220614	100

Ovid MEDLINE(R) and Epub Ahead of Print, In-Process, In-Data-Review & Other Non-Indexed Citations, Daily and Versions <1946 to June 13, 2022>

1	Advance Care Planning/	3861

2	(advance adj (care or health or medical) adj plan*).mp.	5714

3	((advance or anticipatory) adj (plan or plans or planning)).tw.	440

4	Advance Care Plan*.tw.	3900

5	(advance adj (directive* or decision*)).tw.	4130

6	anticipatory care plan*.tw.	38

7	Advance Directives/	6430

8	living will*.tw.	1307

9	Right to Die/	4958

10	right to die.tw.	946

11	ACP.tw.	10606

12	((end of life or EOL) adj5 (care or discuss* or decision* or plan or plans or planning or preference*)).tw.	17847

13	or/1-12	40931

14	Health Personnel/	57932

15	Physicians/	98182

16	Nurses/	43761

17	Nursing assistants/ or Nurses Aides/	4354

18	(nurse* or health care assistant or doctor*).tw.	429704

19	(allied health or doctor* or nurse or nurses or physician* or nursing staff or nursing personnel or therapist* or physiotherapist* or health care professional*).tw.	875761

20	((health or medical) adj (staff or worker* or professional or personnel or provider or practitioner)).tw.	64992

21	or/14-20	1025964

22	Education/	21497

23	(teach* or educat* or learn* or instruct* or train*).tw.	1756417

24	Learning/	75513

25	or/22-24	1778866

26	exp nursing homes/	43082

27	Residential Facilities/	5704

28	Assisted Living Facilities/	1557

29	Long-Term Care/	27736

30	nursing home*1.tw.	33444

31	care home*1.tw.	4895

32	((elderly or old age) adj2 home*1).tw.	2302

33	assisted living facilit*.tw.	840

34	((nursing or residential) adj (home*1 or facilit*)).tw.	39014

35	(home*1 for the aged or home*1 for the elderly or home*1 for older adult*).tw.	3240

36	residential aged care.tw.	1450

37	("frail elderly" adj2 (facilit* or home or homes)).tw.	67

38	(residential adj (care or facilit* or setting*)).tw.	5794

39	or/26-38	94094

40	13 and 21 and 25 and 39	342

41	limit 40 to dt=20180301-20220614	115

APA PsycInfo <1806 to June Week 1 2022>

Added subject headings Advance Directives/ or Decision making/ and Nursing Homes/ or Nursing Home/

1	Advance Directives/ or Decision making/	83696

2	advance care plan*.tw.	1453

3	(advance adj (care or health or medical) adj plan*).mp.	1694

4	(advance adj (directive* or decision*)).tw.	1717

5	anticipatory care plan*.tw.	5

6	living will*.tw.	413

7	right to die.tw.	271

8	ACP.tw.	779

9	((end of life or EOL) adj5 (care or discuss* or decision* or plan or plans or planning or preference*)).tw.	7747

10	or/1-9	90878

11	Health Personnel/	19309

12	PHYSICIANS/ or FAMILY PHYSICIANS/	24957

13	NURSES/	30555

14	Allied Health Personnel/	1234

15	(nurse* or health care assistant or doctor*).tw.	115382

16	(allied health or doctor* or nurse or nurses or physician* or nursing staff or nursing personnel or therapist* or physiotherapist* or health care professional*).tw.	265760

17	((health or medical) adj (staff or worker* or professional or personnel or provider or practitioner)).tw.	21170

18	or/11-17	298771

19	CONTINUING EDUCATION/ or EDUCATION/	42443

20	(teach* or educat* or learn* or instruct* or train*).tw.	1338699

21	exp LEARNING/	289573

22	or/19-21	1421983

23	Nursing Homes/ or Nursing Home/	9361

24	Residential Care Institutions/	11158

25	Assisted Living/	813

26	Long Term Care/	5947

27	nursing home*1.tw.	13169

28	care home*1.tw.	2165

29	((elderly or old age) adj2 home*1).tw.	1312

30	assisted living facilit*.tw.	636

31	((nursing or residential) adj (home*1 or facilit*)).tw.	15850

32	(home*1 for the aged or home*1 for the elderly or home*1 for older adult*).tw.	1501

33	residential aged care.tw.	606

34	("frail elderly" adj2 (facilit* or home or homes)).tw.	39

35	(residential adj (care or facilit* or setting*)).tw.	7323

36	or/23-35	35335

37	10 and 18 and 22 and 36	185

38	book.pt.	518175

39	37 not 38	166

40	limit 39 to english language	162

41	limit 40 to up=20180301-20220614	46

| 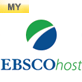CINAHL  Added MH "Advance Care Planning") OR (MH "Decision Making") | Tuesday, June 14, 2022 2:58:35 PM |
| --- | --- |

| **#** | **Query** | **Limiters/Expanders** | **Last Run Via** | **Results** |
| --- | --- | --- | --- | --- |
| S25 | S4 AND S11 AND S15 AND S23 AND EM 201803- | Limiters - Language: English Expanders - Apply equivalent subjects Search modes - Boolean/Phrase | Interface - EBSCOhost Research Databases Search Screen - Advanced Search Database - CINAHL Plus | 83 |
| S24 | S4 AND S11 AND S15 AND S23 | Expanders - Apply equivalent subjects Search modes - Boolean/Phrase | Interface - EBSCOhost Research Databases Search Screen - Advanced Search Database - CINAHL Plus | Display |
| S23 | S16 OR S17 OR S18 OR S19 OR S20 OR S21 OR S22 | Expanders - Apply equivalent subjects Search modes - Boolean/Phrase | Interface - EBSCOhost Research Databases Search Screen - Advanced Search Database - CINAHL Plus | Display |
| S22 | TX "residential care" OR TX "residential facilit*" OR TX "residential setting*" | Expanders - Apply equivalent subjects Search modes - Boolean/Phrase | Interface - EBSCOhost Research Databases Search Screen - Advanced Search Database - CINAHL Plus | Display |
| S21 | TX "home for the aged" OR TX "home for the elderly" | Expanders - Apply equivalent subjects Search modes - Boolean/Phrase | Interface - EBSCOhost Research Databases Search Screen - Advanced Search Database - CINAHL Plus | Display |
| S20 | TX "nursing home*" OR TX "nursing facilit*" OR TX "residential home*" OR TX "residential facilit*" | Expanders - Apply equivalent subjects Search modes - Boolean/Phrase | Interface - EBSCOhost Research Databases Search Screen - Advanced Search Database - CINAHL Plus | Display |
| S19 | AB elderly N3 home* OR AB old age N3 home* | Expanders - Apply equivalent subjects Search modes - Boolean/Phrase | Interface - EBSCOhost Research Databases Search Screen - Advanced Search Database - CINAHL Plus | Display |
| S18 | (MH "Nursing Home Patients") | Expanders - Apply equivalent subjects Search modes - Boolean/Phrase | Interface - EBSCOhost Research Databases Search Screen - Advanced Search Database - CINAHL Plus | Display |
| S17 | (MH "Cancer Care Facilities") OR (MH "Long Term Care") | Expanders - Apply equivalent subjects Search modes - Boolean/Phrase | Interface - EBSCOhost Research Databases Search Screen - Advanced Search Database - CINAHL Plus | Display |
| S16 | (MH "Nursing Homes") | Expanders - Apply equivalent subjects Search modes - Boolean/Phrase | Interface - EBSCOhost Research Databases Search Screen - Advanced Search Database - CINAHL Plus | Display |
| S15 | S12 OR S13 OR S14 | Expanders - Apply equivalent subjects Search modes - Boolean/Phrase | Interface - EBSCOhost Research Databases Search Screen - Advanced Search Database - CINAHL Plus | Display |
| S14 | TX teach* or educat* or learn* or instruct* or train | Expanders - Apply equivalent subjects Search modes - Boolean/Phrase | Interface - EBSCOhost Research Databases Search Screen - Advanced Search Database - CINAHL Plus | Display |
| S13 | (MH "Learning+") | Expanders - Apply equivalent subjects Search modes - Boolean/Phrase | Interface - EBSCOhost Research Databases Search Screen - Advanced Search Database - CINAHL Plus | Display |
| S12 | (MH "Education") | Expanders - Apply equivalent subjects Search modes - Boolean/Phrase | Interface - EBSCOhost Research Databases Search Screen - Advanced Search Database - CINAHL Plus | Display |
| S11 | S5 OR S6 OR S7 OR S8 OR S9 OR S10 | Expanders - Apply equivalent subjects Search modes - Boolean/Phrase | Interface - EBSCOhost Research Databases Search Screen - Advanced Search Database - CINAHL Plus | Display |
| S10 | AB ( "allied health" or "health care assistant" or doctor* or nurse or nurses ) OR AB ( physician* or "nursing staff" or therapist* ) OR AB ( physiotherapist* or practitioner* or GP* ) | Expanders - Apply equivalent subjects Search modes - Boolean/Phrase | Interface - EBSCOhost Research Databases Search Screen - Advanced Search Database - CINAHL Plus | Display |
| S9 | TX "health care professionals" or doctor or nurse of "general practitioner" or "health care worker" | Expanders - Apply equivalent subjects Search modes - Boolean/Phrase | Interface - EBSCOhost Research Databases Search Screen - Advanced Search Database - CINAHL Plus | Display |
| S8 | (MH "Multidisciplinary Care Team") | Expanders - Apply equivalent subjects Search modes - Boolean/Phrase | Interface - EBSCOhost Research Databases Search Screen - Advanced Search Database - CINAHL Plus | Display |
| S7 | (MH "Physicians") OR (MH "Physician Assistants") OR (MH "Geriatricians") | Expanders - Apply equivalent subjects Search modes - Boolean/Phrase | Interface - EBSCOhost Research Databases Search Screen - Advanced Search Database - CINAHL Plus | Display |
| S6 | (MH "Nurses") OR (MH "Practical Nurses") | Expanders - Apply equivalent subjects Search modes - Boolean/Phrase | Interface - EBSCOhost Research Databases Search Screen - Advanced Search Database - CINAHL Plus | Display |
| S5 | (MH "Allied Health Personnel") OR (MH "Health Personnel") | Expanders - Apply equivalent subjects Search modes - Boolean/Phrase | Interface - EBSCOhost Research Databases Search Screen - Advanced Search Database - CINAHL Plus | Display |
| S4 | S1 OR S2 OR S3 | Expanders - Apply equivalent subjects Search modes - Boolean/Phrase | Interface - EBSCOhost Research Databases Search Screen - Advanced Search Database - CINAHL Plus | Display |
| S3 | TX "advance care plan*" OR AB "advance directive*" OR AB "advance decision*" OR AB “anticipatory care plan” OR AB "living well" OR AB "Right to Die" OR AB ACP | Expanders - Apply equivalent subjects Search modes - Boolean/Phrase | Interface - EBSCOhost Research Databases Search Screen - Advanced Search Database - CINAHL Plus | Display |
| S2 | TX "advance care" N3 plan OR TX "advance health" N3 plan OR TX "advance medical" N3 plan | Expanders - Apply equivalent subjects Search modes - Boolean/Phrase | Interface - EBSCOhost Research Databases Search Screen - Advanced Search Database - CINAHL Plus | Display |
| S1 | (MH "Advance Care Planning") OR (MH "Decision Making") | Expanders - Apply equivalent subjects Search modes - Boolean/Phrase | Interface - EBSCOhost Research Databases Search Screen - Advanced Search Database - CINAHL Plus | Display |

**Supplementary Searches – searched 20/6/22**

**Journals**

- American Journal of Hospice and Palliative Medicine
- BMJ Supportive and Palliative care
- Journal of Hospice and palliative nursing
- International Journal of Palliative
- Palliative Medicine

**Websites**

- The National Council for Palliative Care (NCPC): <http://www.professionalpalliativehub.com/resource-centre/national-council-palliative-care>
- Association for Palliative Medicine of Great Britain and Ireland: <https://apmonline.org/>
- Ehospice: <https://ehospice.com/>
- European Association for Palliative Care: <https://www.eapcnet.eu/>
- Gold standards framework: <https://www.goldstandardsframework.org.uk/>
