## Supplementary figures and images for "The Effectiveness of Advance Care Planning Training for Care Home Staff: a Systematic Review"

### Supplementary File 2

**Figure 2:** Ideas Webbing Process

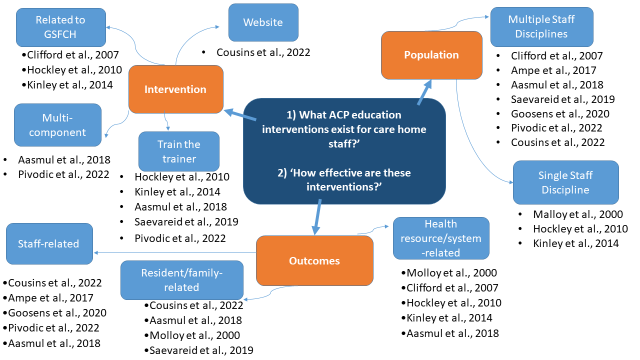
